# Bicalutamide does not raise transaminases in comparison to alternative anti-androgen regimens among transfeminine adolescents and young adults: a retrospective cohort study

**DOI:** 10.1101/2024.02.21.24302999

**Authors:** Katie Burgener, Brian DeBosch, Jinli Wang, Christopher Lewis, Cynthia Herrick

## Abstract

**Background:** Bicalutamide is a potential anti-androgen for transgender individuals with feminizing embodiment goals, but use has been limited because of hepatotoxicity in cisgender men with prostate cancer. This study compared transaminase changes in transfeminine adolescents and young adults (AYA) using low-dose bicalutamide with individuals using other methods of androgen blockade.

**Methods:** A retrospective analysis was conducted using electronic health record data for patients starting gender affirming hormone therapy with at least 10 months of follow-up data between 2015 and 2023. Linear mixed models compared change in ALT and AST from baseline and maximum ALT and AST values in bicalutamide and comparison groups. Secondary outcomes included % individuals with ALT and AST elevation more than 1, 2, or 3 times the upper limit of normal (ULN) (Fisher’s exact test), standardized mean estradiol dose by group (t test), and Tanner staging of breast tissue by group (Fisher’s exact test).

**Results:** Eighty-four transfeminine AYA (median age 18) taking bicalutamide were compared to 69 transfeminine AYA (median age 19) taking GnRH agonists, spironolactone or no agent in addition to estradiol. In linear mixed models adjusted for baseline age, BMI, baseline ALT or AST, and alcohol use, there was no difference in delta or maximum ALT or AST in bicalutamide and comparison groups. No individuals had an AST or ALT level > 3x ULN. Estradiol doses and Tanner stages were similar between groups in a subgroup analysis of individuals receiving pediatric care.

**Conclusion:** Bicalutamide was not associated with significant change in transaminases as compared with other anti-androgen regimens over one year. Bicalutamide appears to be a safe anti-androgen for transfeminine individuals at low dose with close monitoring and deserves further study.

## Introduction

Approximately 1.8% of adolescents in the United States identify as transgender, or having a gender identity that is different from the sex they were assigned at birth, according to the Centers for Disease Control and Prevention (CDC) Youth Risk Behavior Surveillance System (Johns et al, 2017). This estimate of gender diversity in youth has increased from previous estimates based on adult population sampling.

Individuals who identify as transgender have various options available to align physical attributes with their gender identity rather than their birth-assigned sex. These options depend on age, pubertal status, and other factors. For individuals assigned male at birth (AMAB), who are seeking feminine embodiment goals (transfeminine), medical transition options include estradiol with or without an anti-androgen. Anti-androgens currently recommended by the World Professional Association of Transgender Health (WPATH) and the Endocrine Society include GnRH agonists (leuprolide, histrelin) or spironolactone (Coleman et al., 2022; Hembree et al, 2017). In the US, regimen selection depends on a variety of factors, including individual preferences, comorbidities, and insurance coverage.

Bicalutamide is an anti-androgen that blocks the binding of dihydrotestosterone and testosterone to the androgen receptor with subsequent increased aromatization of testosterone to estradiol. Historically, bicalutamide, and other members of the same drug class, have been used in the treatment of prostate cancer. A known side effect of the medication in this population is gynecomastia (Wirth et al., 2004). In AMAB transfeminine individuals, blocking testosterone action and promoting breast development could be a desired effect depending on the individual patient’s goals.

Additionally, given the high cost of GnRH agonists, particularly for individuals with high deductible health plans, and the difficulty obtaining insurance coverage for these agents for gender affirmation, additional anti-androgen options are needed. Spironolactone is the currently recommended oral alternative, although this medication is associated with some adverse effects, including hyperkalemia. This side effect is higher risk in individuals >45 years of age or with underlying renal disease (Hayes et al., 2022). It may also be associated with polyuria, orthostasis, and drowsiness (Patibandla et al., 2023). Lastly, there is some concern for premature breast bud fusion with spironolactone, although this concern requires further investigation (Wierckx et al., 2014).

Bicalutamide is an alternative anti-androgen, but it has not been used routinely because of concern for hepatotoxicity. This is a rare, but potentially serious side effect and has been reported in several case reports and studies when bicalutamide is used for prostate cancer (Blackledge et al., 1997; Dawson et al., 1997; Hussain et al., 2014; Kolvenbag and Blackledge, 1996; McLeod et al., 2006). The degree of hepatotoxicity in these studies ranged from transient liver enzyme elevation to liver failure and death. However, the hepatotoxicity risk is likely correlated with dose, age, and other comorbidities. When it occurs, it is typically within the first three months on therapy. Whereas bicalutamide doses for prostate cancer reach 150 mg daily, doses used in the care of AMAB transfeminine individuals are much lower (25-50 mg daily). Additionally, the risk for hepatotoxicity is substantially higher with other drugs in the class (flutamide) (Bruni et al., 2012).

There is precedent for using bicalutamide in disease states beyond prostate cancer. Bicalutamide has been used in rare forms of precocious puberty in cisgender boys (Lenz et al., 2010; Reiter et al., 2010; Tessaris et al., 2012). It has also been studied in cisgender women with female pattern hair loss and cisgender women with hirsutism in PCOS (Ismail et al., 2020; Moretti et al., 2018). Two studies from a single group examine the use of bicalutamide for adolescent gender affirming care. The first retrospective study reported on 23 individuals on bicalutamide, without a comparison group, finding no AST or ALT elevation on a single measurement among individuals prescribed bicalutamide. All individuals prescribed bicalutamide monotherapy experienced some degree of breast development (Neyman et al., 2019). This group also published a recent study that examined outcomes of 40 individuals on bicalutamide, with no comparison group, finding no liver toxicity (Fuqua et al, 2023). Another small study followed 5 transfeminine individuals prescribed bicalutamide (25mg initially, uptitrated to 50 mg), from 3-12 months. The study reported no major side effects from any medication administered, but it is unclear if liver function testing was followed (Karakılıç Özturan et al., 2023). There is a single current report in the literature of proposed bicalutamide-induced hepatotoxicity in a transfeminine adolescent on bicalutamide 50 mg daily that occurred within three months on therapy (Wilde et al., 2024). Given the potential benefits of bicalutamide, it was incorporated as an option for AMAB transfeminine adolescents and young adults (AYA) seeking gender affirming care at our center beginning in 2017. Patients and their guardians were carefully counselled about the risks and benefits of different anti-androgen options and selected their treatment plan through shared decision making with the center’s endocrinologist.

To date, no studies specifically compare AST and ALT over time among individuals taking bicalutamide and individuals receiving other feminizing therapies. Further, no studies compare estradiol dose among individuals on bicalutamide and individuals on other anti-androgens or regimens without an anti-androgen. Given the paucity of literature on bicalutamide use in gender affirmation, and the proposed safety concerns based on data from older cisgender men with prostate cancer, the primary objective of this study was to identify any significant differences in transaminase levels over one year among individuals on bicalutamide compared with individuals on other feminizing regimens.

## Methods

A retrospective cohort study was conducted utilizing data extracted from the electronic health record. AMAB transfeminine individuals ≤ age 30 newly starting anti-androgen and feminizing hormone therapy with our transgender care team were eligible for inclusion. Depending upon their age at presentation, some individuals were on bicalutamide or GnRH agonist alone for a variable length of time prior to estradiol initiation. Additional inclusion criteria were: on anti-androgen and/or estradiol for at least 1 year with at least 10 months of laboratory data after medication start from January 1, 2015 to February 1, 2023 and prescription of bicalutamide, GnRH agonist or spironolactone and estradiol or estradiol alone. Individuals were excluded if they were assigned female at birth (AFAB) or if they were AMAB not seeking feminizing therapy. Additional exclusion criteria were: absence of baseline or follow-up lab data to at least 10 months after medication start, change in anti-androgen therapy during the follow-up period, or initiation of puberty blockers or gender-affirming hormone therapy (GAHT) at an outside health facility prior to transferring care to our clinic. If an individual had ALT and AST data prior to 10 months post medication start and not again until after 1 year, the first values after 1 year post medication start were utilized. Demographic information (age, gender identity, race, ethnicity and insurance status) and medical and social history relevant to liver dysfunction, (obesity, alcohol or other substance use) were collected. Additionally, exam data (BMI, blood pressure and Tanner staging if documented), and laboratory data (ALT, AST, albumin, estrogen and testosterone levels, prolactin, and lipid panel) were extracted from the medical record. Individuals were assigned a study number and all data were stored in a de-identified REDCap database.

Individuals were included in the bicalutamide group if they remained on bicalutamide for at least one year and on no other anti-androgen in conjunction with estradiol during that follow-up period. Individuals were included in the comparison group if they were on a GnRH agonist, spironolactone, or no anti-androgen in conjunction with estradiol during the follow-up. It was estimated that a sample size of 64 individuals per group would provide 80% power to detect a mean group difference in ALT and AST of 0.5 standard deviation units.

Baseline values in the bicalutamide and comparison groups were compared using t tests, Mann Whitney U test, chi square, or Fisher’s exact test depending on variable type and distribution. ALT and AST differences between groups throughout the follow-up period were assessed in multiple ways. For the primary outcomes of ALT or AST change from baseline and maximum, multivariable linear mixed-effect models with participant as a random effect were used to estimate the adjusted group means and mean differences between groups. Three covariates that were significantly different between groups (baseline ALT or AST, age, and BMI), were included in the models.

The model was also adjusted for reported alcohol use as it could have a potentially important effect on liver function. Additionally, the percentage of individuals in the bicalutamide and comparison groups with an ALT or AST value after baseline that was more than the upper limit of normal (ULN), 2x ULN, and 3x ULN was determined. These outcomes were compared by group utilizing a chi square or Fisher’s exact test as appropriate.

To compare maximal estradiol dose required over the follow-up period, given that different estradiol routes were utilized, we calculated a mean standardized dose for each route of administration (sublingual, injection, transdermal patch) and then utilized the mean standardized variable to compare doses across routes. A t test was used to compare the mean standardized maximum estradiol dose between bicalutamide and comparison groups. A Fisher’s exact test was used to compare maximal Tanner stage (for those in whom it was documented) over the one-year follow-up between groups.

The study was approved by the Washington University Human Research Protection Office on December 15, 2021. Statistical analysis was performed using SAS v 9.4 (Cary, NC).

## Results

Three hundred and seventy-eight transfeminine AMAB individuals ≤ 30 years of age that had received gender affirming hormone therapy in our transgender center were initially identified using the Slicer Dicer tool on our Epic electronic health record. Of these individuals, 123 were excluded for either lack of baseline labs, inadequate monitoring labs over year of follow-up, or treatment for less than one year at the time of data extraction. An additional 90 individuals were excluded because they had initiated GAHT prior to presentation to our transgender center. Lastly, one individual changed anti-androgen therapy and was excluded. One hundred and sixty-four individuals had data extracted to the REDCap database. After data quality assessment, an additional 11 individuals were excluded (3 for initiation of GAHT prior to establishing with the center, 4 for inadequate baseline labs, and 4 for switching anti-androgen regimens during the one-year follow-up). Individuals who switched anti-androgen therapy did not do so because of AST or ALT abnormalities. After these exclusions, 84 individuals in the bicalutamide group and 69 individuals in the comparison group were included in the analyses (Figure 1). The mean (SD) follow-up time from medication start to last ALT and AST extracted in the cohort was 394 (113) days.

**Figure 1.**
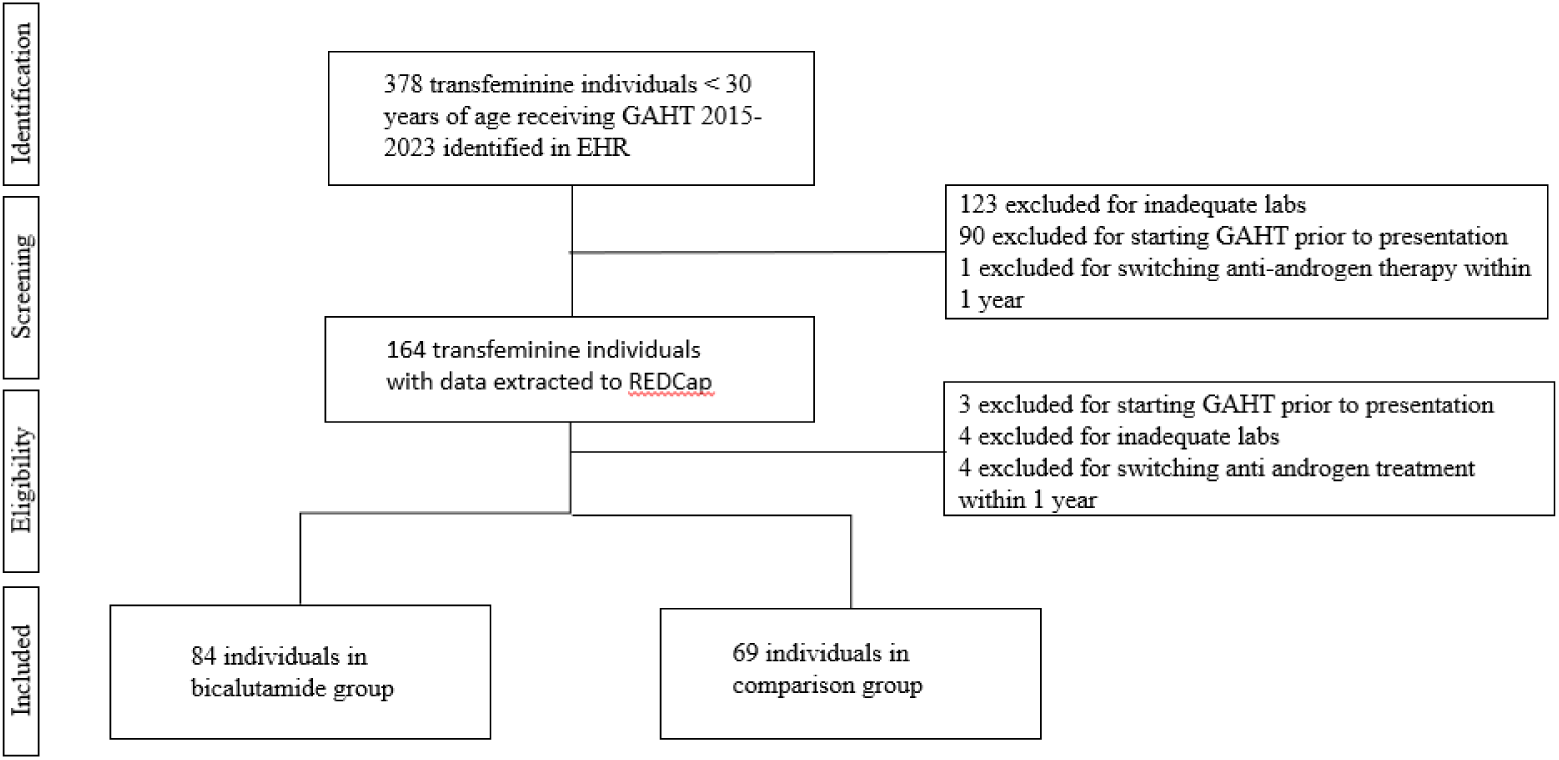
PRISMA diagram for AMAB transfeminine individual retrospective cohort definition

Baseline demographic, medical history, exam, and lab data are shown in Table1. Median age of the total population was 18 years (IQR 16, 20). Approximately 91% identified as binary transgender females while approximately 9% identified as non-binary, genderqueer or agender seeking feminine embodiment goals. The majority of individuals identified as white (79.7%), whereas 11.8% identified as Black, 2% as Asian, 4.6% as Hispanic, and 2% as another or multiple races/ethnicities. The majority (78.4%) had private insurance, 17% had Medicaid, 2.6% had other government insurance, and 2% were uninsured. Individuals in the bicalutamide group were younger than the comparison group (median age 18 (IQR 16,18) v. 19 (IQR 17,24) years, p < 0.0001). There were no statistically significant differences between groups for gender identity, race/ethnicity, or insurance carrier. There were also no significant differences between groups in prevalence of type 1 or type 2 diabetes mellitus, hyperlipidemia, hypertension, HIV, alcohol use, or other major medical problems that may influence baseline liver function testing. Median BMI was significantly different between groups (bicalutamide, 21.6 kg/m^2^ (IQR 18.9, 25.3) v. comparison, 25.5 kg/m^2^ (IQR 21.0, 30.4), p=0.001). Baseline median ALT and AST were also significantly different between groups [median ALT: bicalutamide,14 U/L (IQR 11, 22) v comparison, 21 U/L (IQR 13, 32), p=0.002; median AST bicalutamide, 19 U/L (IQR 16, 23) v comparison, 21 U/L (IQR 18, 29), p=0.008].

Tables 2 and 3 summarize linear mixed models with outcomes of change in ALT and AST over baseline and maximum ALT and AST levels during the follow-up period. Adjusted ALT declined in both bicalutamide [-3.8 U/L (95% CI -7.1, -0.5)] and comparison groups [-7.3 U/L (95% CI -11.3, -3.2] over the follow-up period. The adjusted maximum ALT levels in the bicalutamide and comparison groups were not elevated in either group [bicalutamide, 22.1 U/L (95% CI 18.8, 25.4) v. comparison, 18.7 U/L (95% CI 14.6, 22.8)]. There was no significant between-group difference in adjusted delta ALT or maximum ALT (3.4 U/L, 95% CI -0.6, 7.5; p= 0.09). The adjusted AST change from baseline in the bicalutamide and comparison groups was -1.6 U/L (95% CI -3.3, 0.1) and -2.7 U/L (95% CI -4.8, -0.5.), respectively. The adjusted maximum AST levels in the bicalutamide and comparison groups were 20.1 U/L (95% CI 18.4, 21.8) and 19.0 U/L (95% CI 16.9, 21.2), respectively. No significant differences in delta AST or maximum AST were noted between the bicalutamide and comparison group (1.1 U/L, 95% CI -1.1, 3.2, p= 0.32).

**Table 1:** Baseline characteristics of AMAB transfeminine retrospective cohort by treatment group

| Variable | Total Population<br>n=153 | Bicalutamide<br>n=84 | Comparison<br>n=69 | P-value |
| --- | --- | --- | --- | --- |
| Age (at first encounter), Median (IQR) | 18 (16, 20)<br>Range: 12-30 | 18 (16, 18) | 19 (17, 24) | <0.0001<sup>a</sup> |
| Gender identity, n (%) |  |  |  | 0.86 |
| Binary transgender female | 139 (90.8%) | 76 (90.5%) | 63 (91.3%) |  |
| Another gender identity with feminine embodiment goals (nonbinary, agender, genderqueer) | 14 (9.2%) | 8 (9.5%) | 6 (8.7%) |  |
| Race/Ethnicity, n (%) |  |  |  | 0.59<sup>b</sup> |
| Asian, non-Hispanic | 3 (2%) | 1 (1.2%) | 2 (2.9%) |  |
| Black, non-Hispanic | 18 (11.8%) | 7 (8.3%) | 11 (15.9%) |  |
| White, non-Hispanic | 122 (79.7%) | 70 (83.3%) | 52 (75.4%) |  |
| Hispanic | 7 (4.6%) | 4 (4.8%) | 3 (4.3%) |  |
| Another race/ethnicity or multiple race/ethnicities | 3 (2%) | 2 (2.4%) | 1 (1.4%) |  |
| Primary insurance, n(%) |  |  |  | 0.10<sup>b</sup> |
| Private | 120 (78.4%) | 64 (76.2%) | 56 (81.2%) |  |
| Medicaid | 26 (17%) | 18 (21.4%) | 8 (11.6%) |  |
| Other government insurance | 4 (2.6%) | 2 (2.4%) | 2 (2.9%) |  |
| Uninsured | 3 (2%) | 0 (0%) | 3 (4.3%) |  |
| Medical History, n (%) |  |  |  |  |
| No medical history | 35 (22.9%) | 19 (22.6%) | 16 (23.2%) | 0.93 |
| Type 1 diabetes | 1 (0.7%) | 1 (1.2%) | 0 (0%) | >0.99<sup>b</sup> |
| Type 2 diabetes | 0 (0%) | 0 (0%) | 0 (0%) | NA |
| Hyperlipidemia | 2 (1.3%) | 2 (2.4%) | 0 (0%) | 0.50<sup>b</sup> |
| Hypertension | 2 (1.3%) | 1 (1.2%) | 1 (1.4%) | >0.99<sup>b</sup> |
| Liver disease | 0 (0%) | 0 (0%) | 0 (0%) | NA |
| HIV | 4 (2.6%) | 1 (1.2%) | 3 (4.3%) | 0.33<sup>b</sup> |
| Medical problem not listed | 106 (69.3%) | 61 (72.6%) | 45 (65.2%) | 0.32 |
| Reported alcohol use, n (%) | 13 (8.5%) | 10 (11.9%) | 3 (4.3%) | 0.10 |
| Reported substance use, n (%) | 18 (11.8%) | 13 (15.5%) | 5 (7.2%) | 0.12 |
| BMI, Median (IQR) | 22.4 (19.7, 28.3),<br>n=152 | 21.6 (18.9, 25.3) | 25.5 (21.0, 30.4),<br>n=68 | 0.001<sup>a</sup> |
| Systolic blood pressure, mean (SD) | 125.2 (13.0), n=149 | 122 (11.2), n=82 | 129.2 (14), n=67 | 0.0006 |
| Diastolic blood pressure, mean (SD) | 74 (9.7), n=149 | 70.4 (8.3), n=82 | 78.5 (9.5), n=67 | <.0001 |
| Baseline ALT |  |  |  |  |
| ALT (Units/L), median (IQR) | 16 (12, 28) | 14 (11, 22) | 21 (13, 32) | 0.002<sup>a</sup> |
| ALT > 2x ULN | 2 (1.3%) | 0 (0%) | 2 (2.9%) | 0.20<sup>b</sup> |
| ALT > 3x ULN | 0 (0%) | 0 (0%) | 0 (0%) | NA |
| Baseline AST | n=152 |  | n=68 |  |
| AST (Units/L), median (IQR) | 19 (16, 24) | 19 (16, 23) | 21 (18, 29) | 0.008<sup>a</sup> |
| AST > 2x ULN | 0 (0%) | 0 (0%) | 0 (0%) | NA |
| AST > 3x ULN | 0 (0%) | 0 (0%) | 0 (0%) | NA |

Abbreviations: n= sample size; SD= standard deviation; IQR= Interquartile Range; NA= not applicable
When less than the entire cohort provided data, sample size (n) or the denominator is also reported
<sup>a</sup> P-value is from Wilcoxon Rank Sum Test due to the small sample size and skewness of data. Median (IQR) are reported.
<sup>b</sup> P-value are from Fishers' exact test because $\geq 25\%$ of the cells have expected counts less than 5

**Table 2.**
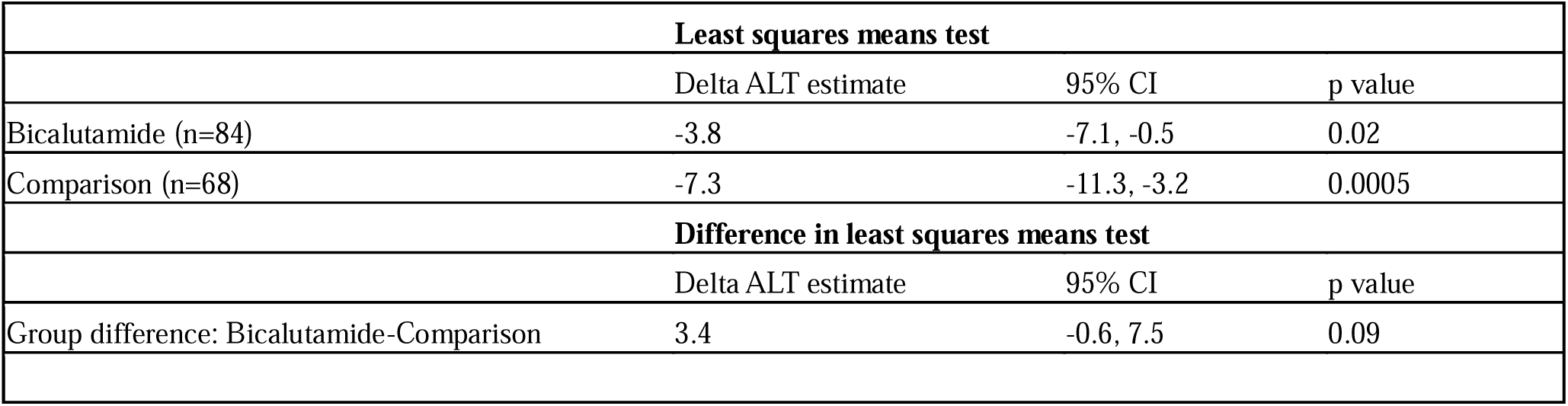
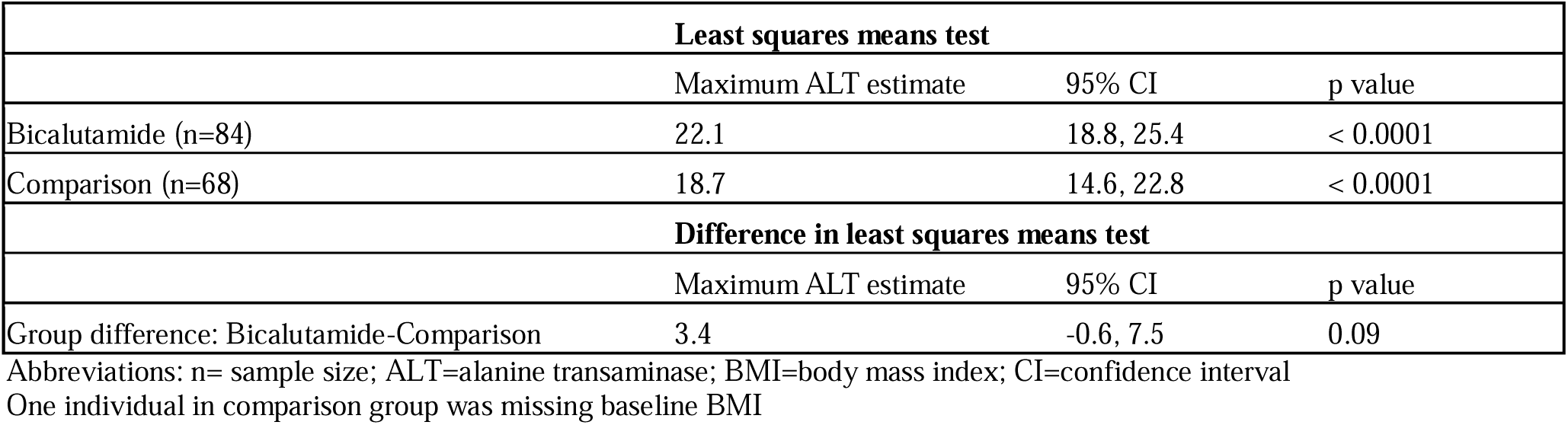
a: Linear mixed model examining fixed effect of medication group on delta ALT, adjusting for age, BMI, alcohol use, and baseline ALT (n=152) b: Linear mixed model examining fixed effect of medication group on maximum ALT, adjusting for age, BMI, alcohol use, and baseline ALT (n=152)

**Table 3.**
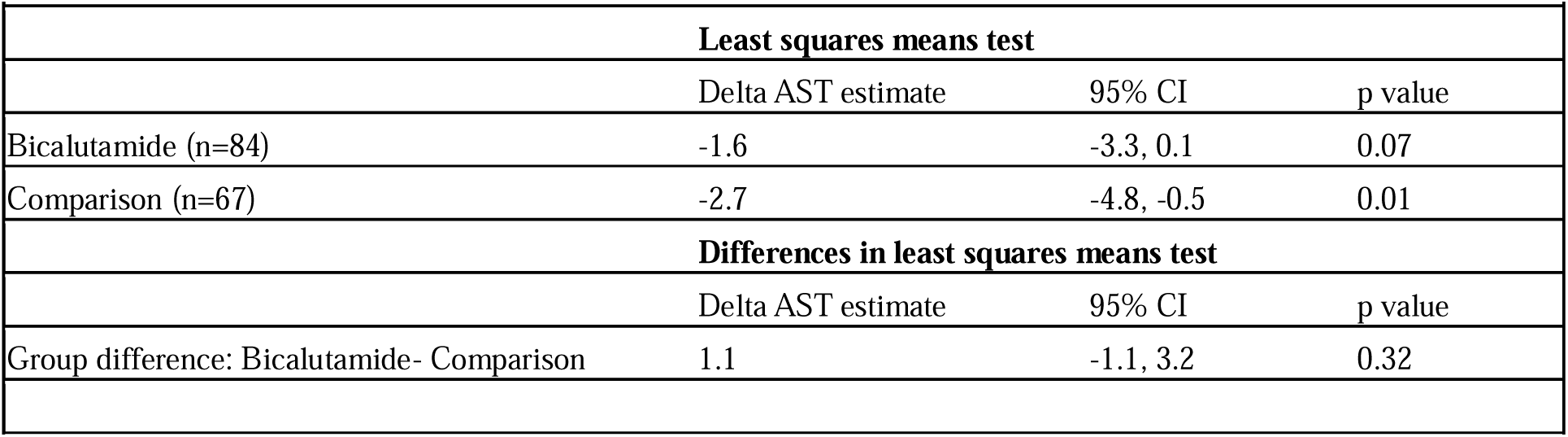
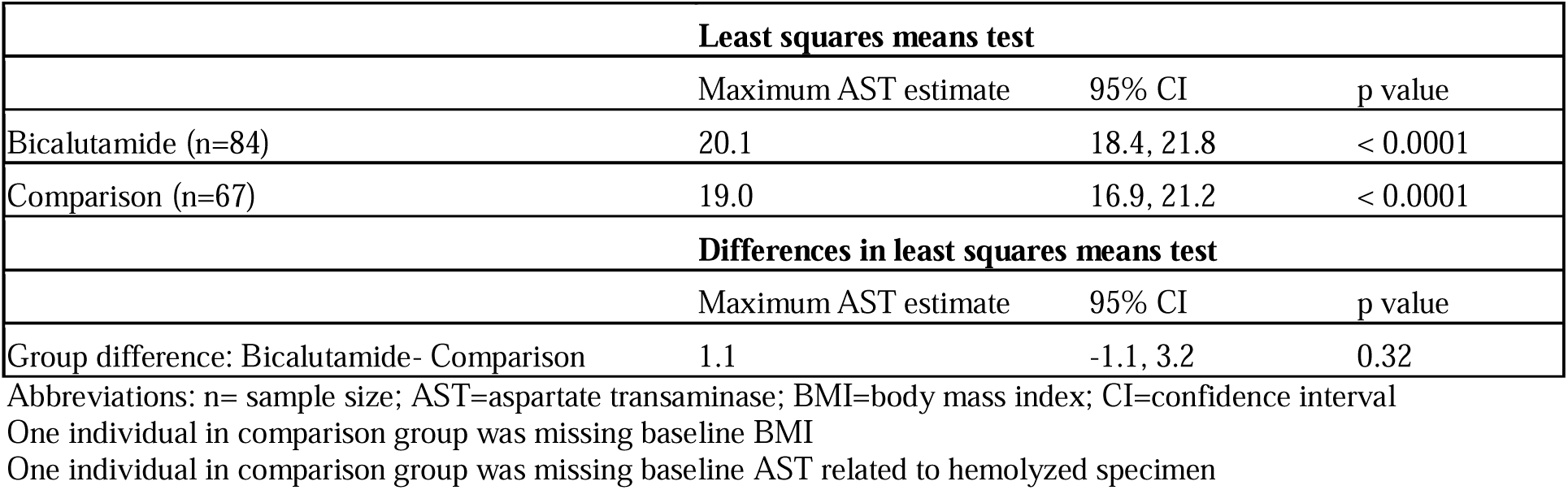
a: Linear mixed model examining fixed effect on medication group on delta AST, adjusting for age, BMI, alcohol use, and baseline AST (n=151) b: Linear mixed model examining fixed effect of medication group on maximum AST, adjusting for age, BMI, alcohol use, and baseline AST (n=151)

One individual in comparison group was missing baseline BMI
|  |  |  |  |
| --- | --- | --- | --- |
| Table 3a: Linear mixed model examining fixed effect on medication group on delta AST, adjusting for age, BMI, alcohol use, and baseline AST (n=151) |  |  |  |
| **Least squares means test** |  |  |  |
|  | Delta AST estimate | 95% CI | p value |
| Bicalutamide (n=84) | -1.6 | -3.3, 0.1 | 0.07 |
| Comparison (n=67) | -2.7 | -4.8, -0.5 | 0.01 |
| **Differences in least squares means test** |  |  |  |
|  | Delta AST estimate | 95% CI | p value |
| Group difference: Bicalutamide- Comparison | 1.1 | -1.1, 3.2 | 0.32 |
| Table 3b: Linear mixed model examining fixed effect of medication group on maximum AST, adjusting for age, BMI, alcohol use, and baseline AST (n=151) |  |  |  |
| **Least squares means test** |  |  |  |
|  | Maximum AST estimate | 95% CI | p value |
| Bicalutamide (n=84) | 20.1 | 18.4, 21.8 | < 0.0001 |
| Comparison (n=67) | 19.0 | 16.9, 21.2 | < 0.0001 |
| **Differences in least squares means test** |  |  |  |
|  | Maximum AST estimate | 95% CI | p value |
| Group difference: Bicalutamide- Comparison | 1.1 | -1.1, 3.2 | 0.32 |

Abbreviations: n= sample size; AST=aspartate transaminase; BMI=body mass index; CI=confidence interval
One individual in comparison group was missing baseline AST related to hemolyzed specimen

Table 4 demonstrates individuals in the bicalutamide and comparison groups with any ALT or AST levels >ULN, > 2x ULN, and > 3x ULN during the 1-year follow-up period. There were no significant differences in the % of individuals with any ALT > ULN in each group (16.7% bicalutamide v. 11.6% comparison; p= 0.37). There was a significant difference in percentage of individuals with any AST > ULN (10.7% bicalutamide v. 1.5% comparison; p= 0.02). However, few individuals had AST or ALT > 2x ULN and no individuals in the bicalutamide or comparison groups had any ALT or AST > 3x ULN (typically considered clinically significant transaminitis). For individuals with either baseline elevations in ALT or AST or individuals who developed elevations during the follow up period, 64% of those in the bicalutamide group and 63% of those in the comparison group had normalization of their ALT levels over 1 year. Similarly, 56% in the bicalutamide group and 100% in the comparison group had normalization of their AST by the 1-year follow up laboratory assessment (Supplemental table 1).

**Table 4:** Comparison of maximum ALT and AST elevations in relation to upper limit of normal by medication group (n=153)

One individual in comparison group was missing baseline BMI One individual in comparison group was missing baseline AST related to hemolyzed specimen
|  |  |  |  |  |
| --- | --- | --- | --- | --- |
|  | **ALT n (%)** | **P value** | **AST n (%)** | **P value** |
|  | **> ULN** |  | **> ULN** |  |
| Bicalutamide | 14 (16.7) | 0.37 | 9 (10.7) | 0.02 |
| Comparison | 8 (11.6) |  | 1 (1.5) |  |
|  | **> 2x ULN** |  | **>2x ULN** |  |
| Bicalutamide | 2 (2.4) | > 0.99 | 0 (0) | 0.45 |
| Comparison | 1 (1.5) |  | 1 (1.5) |  |
|  | **> 3x ULN** |  | **> 3x ULN** |  |
| Bicalutamide | 0 (0) |  | 0 (0) |  |
| Comparison | 0 (0) |  | 0 (0) |  |

Abbreviations: n=sample size; ALT=alanine transaminase; AST=aspartate transaminase;
ULN=upper limit of normal
\*P value calculated using chi square or Fisher's exact test

Estrogen dosing and Tanner staging data are shown in tables 5 and 6. The mean standardized maximum daily estradiol dose [mean (SD)] was -0.43 (0.68) in the bicalutamide group v. 0.51 (1.05) in the comparison group, which was statistically significant (p < 0.001) (Table 5a). However, given practice differences in the typical starting estradiol dose and titration among pediatric and adult-trained endocrinologists, a sub-analysis of individuals who were treated only by pediatrics-trained practitioners was completed. In this subgroup, there was no significant difference between standardized maximum daily estradiol dose in the bicalutamide and comparison groups (bicalutamide 0.04 (0.96) v. comparison -0.10 (1.16), p=0.57) (Table 5b). Among individuals seen by pediatric-trained providers, there was also no significant difference in maximum daily estradiol dose for either sublingual or transdermal routes of administration.

**Table 5.**
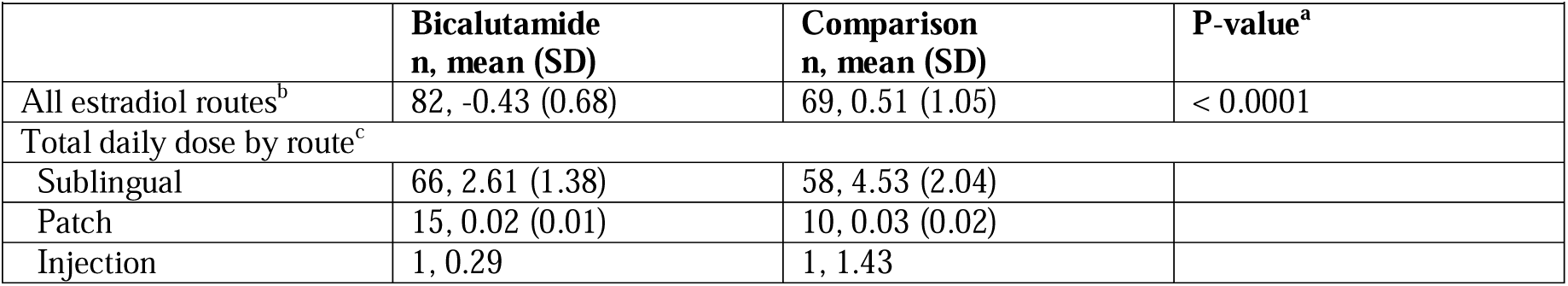
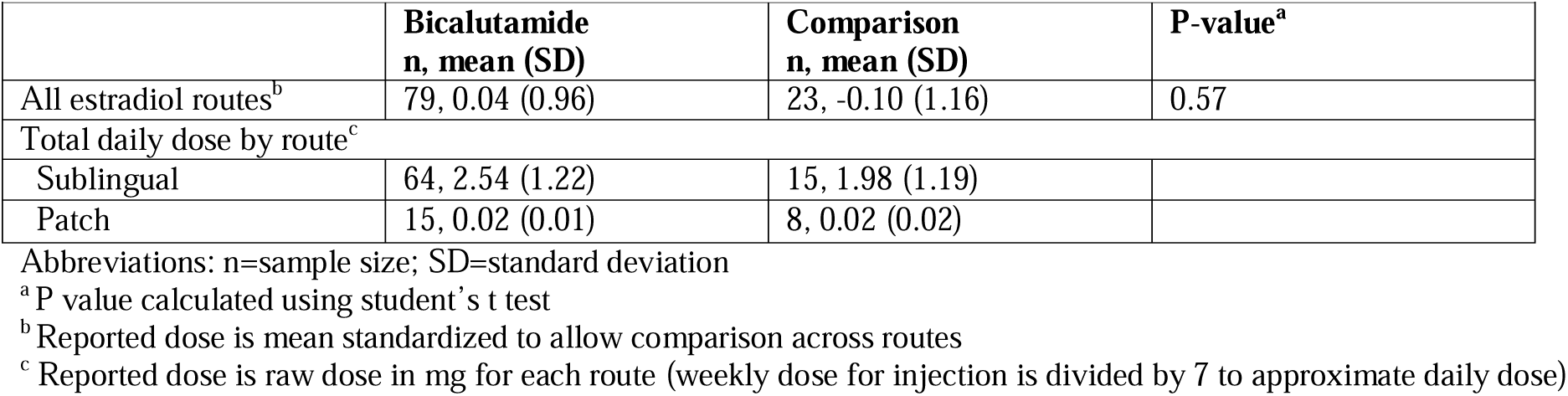
a: Maximum Daily Estradiol Dose in Bicalutamide vs Comparison Groups for Entire Study Population (n=151) b: Maximum Daily Estradiol Dose in Bicalutamide vs Comparison Groups, for Individuals Treated by Pediatric Providers (n= 102)

| Table 5a: Maximum Daily Estradiol Dose in Bicalutamide vs Comparison Groups for Entire Study Population (n=151) |  |  |  |
| --- | --- | --- | --- |
|  | **Bicalutamide**<br>**n, mean (SD)** | **Comparison**<br>**n, mean (SD)** | **P-value<sup>a</sup>** |
| All estradiol routes<sup>b</sup> | 82, -0.43 (0.68) | 69, 0.51 (1.05) | < 0.0001 |
| Total daily dose by route<sup>c</sup> |  |  |  |
| Sublingual | 66, 2.61 (1.38) | 58, 4.53 (2.04) |  |
| Patch | 15, 0.02 (0.01) | 10, 0.03 (0.02) |  |
| Injection | 1, 0.29 | 1, 1.43 |  |
| Table 5b: Maximum Daily Estradiol Dose in Bicalutamide vs Comparison Groups, for Individuals Treated by Pediatric Providers (n= 102) |  |  |  |
|  | **Bicalutamide**<br>**n, mean (SD)** | **Comparison**<br>**n, mean (SD)** | **P-value<sup>a</sup>** |
| All estradiol routes<sup>b</sup> | 79, 0.04 (0.96) | 23, -0.10 (1.16) | 0.57 |
| Total daily dose by route<sup>c</sup> |  |  |  |
| Sublingual | 64, 2.54 (1.22) | 15, 1.98 (1.19) |  |
| Patch | 15, 0.02 (0.01) | 8, 0.02 (0.02) |  |

Abbreviations: n=sample size; SD=standard deviation
<sup>a</sup> P value calculated using student's t test
<sup>b</sup> Reported dose is mean standardized to allow comparison across routes
<sup>c</sup> Reported dose is raw dose in mg for each route (weekly dose for injection is divided by 7 to approximate daily dose)

**Table 6:**
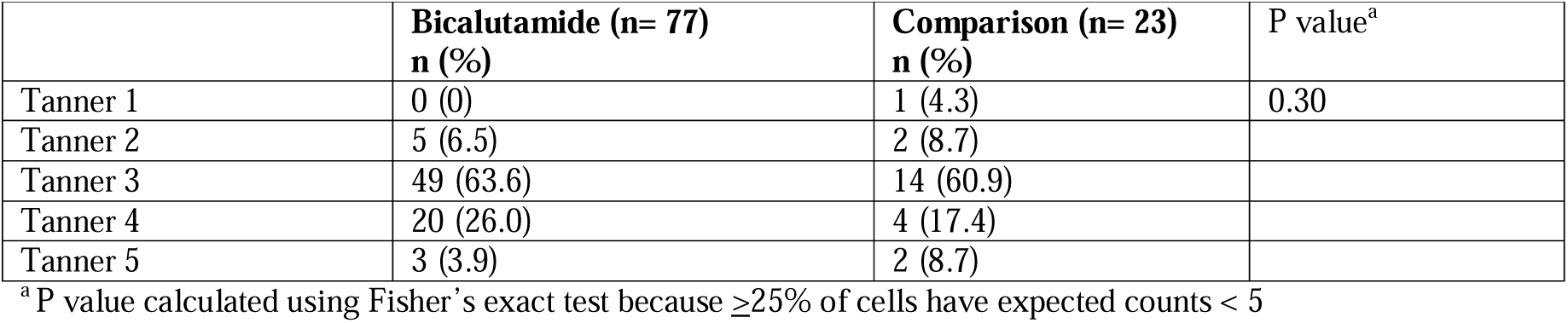
Maximum achieved breast Tanner stage at end of year follow-up by group (n=100)

|  |  |  |  |
| --- | --- | --- | --- |
|  | **Bicalutamide (n= 77)**<br>**n (%)** | **Comparison (n= 23)**<br>**n (%)** | **P value<sup>a</sup>** |
| Tanner 1 | 0 (0) | 1 (4.3) | 0.30 |
| Tanner 2 | 5 (6.5) | 2 (8.7) |  |
| Tanner 3 | 49 (63.6) | 14 (60.9) |  |
| Tanner 4 | 20 (26.0) | 4 (17.4) |  |
| Tanner 5 | 3 (3.9) | 2 (8.7) |  |

<sup>a</sup> P value calculated using Fisher's exact test because $\geq 25\%$ of cells have expected counts < 5

Tanner staging data were missing among a large number of individuals in the comparison group, again related to differences in practice and documentation between pediatric and adult-trained endocrinologists. A majority of patients in the bicalutamide group had at least Tanner stage 3 breast development after a minimum of 6 months of bicalutamide treatment, and there was no difference in Tanner stage between groups (Table 6).

## Discussion

This retrospective cohort study demonstrates no clinically significant differences in ALT and AST values in transfeminine individuals taking bicalutamide when compared with a group of individuals on GnRH agonists, spironolactone or estradiol alone. There have only been three published studies evaluating the use of bicalutamide in AMAB transfeminine individuals (Fuqua et al., 2023; Karakılıç Özturan et al., 2023; Neyman et al., 2019). Our study is the largest study to date in this population and the first study in AMAB transfeminine individuals to systematically evaluate liver function changes on bicalutamide with respect to a comparison group. Additional exploratory findings from our study suggest that estradiol dose requirements are likely similar on bicalutamide and other anti-androgen regimens after accounting for differential prescribing practices. Individuals on bicalutamide also appear to reach appropriate Tanner stage 3 breast development in the first year on therapy, similar to those on other anti-androgens.

Previous studies evaluating bicalutamide in adult cisgender men being treated for prostate cancer demonstrated rare but occasionally serious elevations in transaminases, notably leading to severe hepatic failure and death in some case reports (Blackledge et al., 1997; Dawson et al., 1997; Hussain et al., 2014; Kolvenbag and Blackledge, 1996; McLeod et al., 2006). Bicalutamide doses used in prostate cancer are up to 150 mg daily. Due to these concerns of liver toxicity, bicalutamide has not been routinely used as an anti-androgen in AMAB transfeminine individuals, despite the much lower doses needed in this population (∼25-50 mg daily). One case report published in 2024 described a transgender female adolescent prescribed bicalutamide 50 mg daily who presented to a hospital with liver toxicity that resolved after stopping bicalutamide (Wilde et al., 2024). This appears to be the first documented case of bicalutamide-induced hepatoxicity in a transgender female. The largest published series of transfeminine adolescents on bicalutamide prior to our study showed no significant ALT or AST abnormalities in 40 individuals (Fuqua et al., 2023).

There are other examples in the literature of studies utilizing bicalutamide for indications outside of prostate cancer, with doses that are closer to those used for gender affirmation, but do not demonstrate major adverse effects from bicalutamide use. Bicalutamide has been used as an adjunct treatment (with minoxidil) for cisgender women with female pattern hair loss (Ismail et al., 2020). A retrospective review of 316 cisgender females, mean 49 years of age (range 15-85), prescribed bicalutamide (dose range 5 mg-50 mg daily; standard dose 10 mg daily) demonstrated reassuring safety profiles in individuals using bicalutamide, with 2.9% having mild elevation in liver transaminases, no greater than 2x ULN (Ismail et al., 2020). Additionally, there was subsequent normalization of transaminases in about half of the patients without a dose reduction. This is consistent with the findings of our study. In a double blind randomized controlled trial of 88 cisgender women, a dose of bicalutamide 50 mg daily was evaluated in combination with oral contraceptives in the treatment of hirsutism in PCOS. No significant changes in ALT and AST were documented in the year the randomized study was conducted (Moretti et al., 2018). Finally, bicalutamide has also been used for the treatment of testotoxicosis (in addition to a 3^rd^ generation aromatase inhibitor) at various dosing regimens. A case report of 2 boys with testotoxicosis described long term use of bicalutamide (around 4 years) with no resultant liver function abnormalities (Lenz et al., 2010). One phase 2 trial evaluating use of anastrozole and bicalutamide documented 1 of the 14 boys developed ALT/AST elevation (which was not described as a clinically serious adverse effect) (Reiter et al., 2010). A case report using the same combination in the treatment of a boy with McCune Albright syndrome also showed no elevation in liver enzymes during his approximately two-year treatment period (Tessaris et al., 2012).

In transfeminine individuals seeking gender affirming hormone therapy, our study demonstrated no difference in change in ALT or AST from baseline, maximum ALT or AST values or clinically significant transaminitis (defined as ALT or AST > 3x ULN) between individuals using bicalutamide (25mg daily) and individuals in a comparison group on GnRH agonists, spironolactone or estradiol alone. In fact, ALT and AST declined in both groups over the study period. Bicalutamide use in this study population was associated with lower to no significant difference in mean standardized estradiol doses with the comparison group and demonstrated promising results for breast development based on limited Tanner staging data. There was one individual in whom bicalutamide was stopped after the follow-up period designated for the study.

This individual developed ALT and AST >2x ULN after an episode of COVID and had a thorough hepatology evaluation. As ALT and AST were never > 3x ULN, it was not recommended that bicalutamide be stopped; however, ultimately a clinical decision was made to stop the medication and ALT and AST normalized. Based on these findings, bicalutamide appears to be a safe anti-androgen adjunct to estradiol therapy in transfeminine individuals. In our experience, it is often covered by insurance carriers and is another oral option for patients seeking an alternative to injectable or implantable GnRH agonists.

This study has several limitations. First, this is a retrospective study with data collected via electronic health record chart review. Data were fully extracted and reviewed by one author (KB). A second reviewer performed quality assurance and data validation on > 50% of charts to mitigate risk of human error in data extraction. Data extractors were not blinded to treatment assignment; however, individuals extracting data and performing quality assurance were not the same individuals who were prescribing the medications. Retrospective data collection is limited by availability in existing records. Some data, particularly Tanner staging, were missing in patient notes.

ALT is an incompletely sensitive marker of liver injury, with one study demonstrating that an elevated ALT can occur in biopsy-proven normal liver, but ALT can also be normal in those with biopsy-proven liver injury (Bedgoni et al., 2005). However, ALT screening is still arguably the best non-invasive test currently available to initially screen for drug-induced hepatocellular liver dysfunction (European Association for the Study of the Liver, 2019). Additionally, while labs were scheduled every three months, some patients did not get labs completed on schedule. Moreover, given that individuals were not prospectively assigned or randomized to bicalutamide or comparison groups, there were differences in some baseline characteristics between groups. These limitations in retrospective study design were mitigated by the use of linear mixed models in statistical analysis. Finally, the study was completed using data from a single center and patients in their teens and twenties, which could limit the generalizability of findings to other centers and older patients.

While our sample size was larger than previous studies examining bicalutamide in transfeminine individuals, the study was powered to detect a difference in AST and ALT of 0.5 standard deviation units. Hence, although the study was not powered to detect a smaller difference in these laboratory values, smaller differences are unlikely to be clinically significant. The small size of the study would also not have detected rare adverse events occurring in less than one percent of the exposed population.

Nonetheless, despite these limitations, this study provides reassuring data as to the safety of low dose bicalutamide as an anti-androgen option in feminizing regimens for AMAB transfeminine individuals. It also provides preliminary data to support additional larger, prospective studies comparing bicalutamide to other anti-androgens (spironolactone), GnRH agonists, or estradiol use alone, examining outcomes such as patient satisfaction with their regimen, liver function abnormalities, estradiol dose and level attainment, fertility, and breast development.

## Data Availability

All data produced in the present study are available upon reasonable request to the authors.

## Acknowledgements

We would like to acknowledge Eric Lei who assisted with data quality control and Michael Wallendorf who provided initial statistical planning assistance and preliminary analysis.

## Author contributions

KB designed the study, performed data collection and quality assurance and wrote and edited the manuscript. BD contributed to study design, provided pediatric hepatology expertise, and reviewed and edited the manuscript. JW performed data analysis and reviewed and edited the manuscript. CL contributed to study conceptualization and design, provided pediatric endocrinology expertise, and reviewed and edited the manuscript. CJH contributed to study conceptualization and design, performed data validation, reviewed and edited the manuscript and supervised all aspects of the study.

## Disclosure statement

The authors report there are no competing interests to declare. Ethical Approval:

All procedures performed in studies involving human participants were in accordance with the ethical standards of the institutional and/or national research committee and with the 1964 Helsinki declaration and its later amendments or comparable ethical standards. For this type of study formal consent is not required.

