## Supplemental Table 1 for "Bicalutamide does not raise transaminases in comparison to alternative anti-androgen regimens among transfeminine adolescents and young adults: a retrospective cohort study"

Supplemental Table 1: Comparison of Rates of Normalization of Abnormal ALT and AST Levels during one-year follow-up among AMAB transfeminine individuals on bicalutamide or other anti-androgen regimens

| **Variable** | **Total Population** | **Bicalutamide** | **Comparison** |
| --- | --- | --- | --- |
| ALT | n=22 | n=14 | n=8 |
| Normalized | 14 (64%) | 9 (64%) | 5 (63%) |
| Remained elevated | 8 (36%) | 5 (36%) | 3 (37%) |
| AST | n=10 | n=9 | n=1 |
| Normalized | 6 (60%) | 5 (56%) | 1 (100%) |
| Remained elevated | 4 (40%) | 4 (44%) | 0 (0%) |
